# Analytical and clinical validation of step counting method in people living with amyotrophic lateral sclerosis

**DOI:** 10.1101/2025.07.28.25332300

**Authors:** Marcin Straczkiewicz, Katherine M. Burke, Kendall T. Carney, Narghes Calcagno, Sravan Mandepudi, Alan Premasiri, Fernando G. Vieira, James D. Berry

## Abstract

**Background:** Accelerometer-based digital measures offer a scalable and low-burden means of quantifying physical function, but existing processing algorithms may not quantify pathological gait correctly. In people living with amyotrophic lateral sclerosis (ALS), where gait patterns are slow, variable, and asymmetric, validated tools to quantify mobility are urgently needed.

**Methods:** We proposed a step-counting algorithm designed for ankle-worn accelerometers that leverage wavelet-based decomposition to quantify heel strikes under heterogeneous gait patterns. We validated this method using five datasets comprised of healthy individuals and those with ALS in controlled and semi-controlled activities, and we performed clinical validation in a free-living cohort of 305 people with ALS. We tested our method for accuracy in detecting steps and recognizing walking activity. Reference labels used for analytical validation were obtained from annotated studies or video-based ground truth. Step counting accuracy was assessed using Bland-Altman analysis while clinical validity was evaluated by comparing step counts to gross motor functioning on the ALS Functional Rating Scale – Revised (ALSFRS-R).

**Results:** Walking recognition was robust across walking conditions and body types; sensitivity ranged from 0.94 to 0.98, and specificity exceeded 0.95 across all evaluated datasets. The mean step counting bias was minimal (e.g., 0.44 steps), and the 95% limits of agreement were narrow (LoA = [−5.90, 5.40]) relative to reference standards, including video-annotated ground truth. Clinical validation indicated substantial differences between groups with various levels of gait impairment, e.g., participants who reported “walks with assist” on the ALSFRS-R accumulated a mean of 1283 (95% CI: 1063, 1503) steps/day, while those reporting “normal” walking covered 3984 (95% CI: 3537, 4432) steps/day.

**Conclusions:** Our study covered analytical and clinical validation of a step-counting method developed for ankle-worn accelerometers and demonstrated its applicability to pathological gait. The method provides accurate quantification of walking activity in controlled and free-living environments, supporting its use as a digital endpoint in ALS research.

## 1. Background

Accelerometer-based step counts have emerged as a valuable indicator of physical functioning in both clinical and epidemiological research, offering a scalable, low-burden approach to monitoring mobility across diverse populations. Gait speed has been referred to as the “sixth vital sign” due to its strong association with functional status, fall risk, hospitalization, and mortality (1–3). In people living with amyotrophic lateral sclerosis (ALS), a progressive neurodegenerative condition characterized by motor neuron loss, functional decline, and ultimately respiratory failure, capturing ambulatory function is particularly critical. Gait impairment is often among the earliest manifestations of ALS (4–6), and progressively worsens, leading to decreased mobility, increased fall risk, and compounding health complications (7–9).

Despite its clinical relevance, mobility assessment in ALS has traditionally relied on infrequent and coarse-grained outcome measures such as the ALS Functional Rating Scale-Revised (ALSFRS-R), which may fail to detect subtle yet meaningful changes in walking behavior (10–13). As therapeutics evolve, there is an urgent need for sensitive, responsive, and ecologically valid outcome measures capable of detecting functional improvements and disease progression (14–16).

Digital health technologies (DHTs), including wearable accelerometers and smartphones, enable continuous, high-frequency assessment of motor function in both clinical and free-living environments (17–20). However, existing step counting algorithms are often developed for healthy populations and may perform poorly in individuals with slower, asymmetric, or variable gait patterns typical to ALS (21,22). Furthermore, the clinical validity of step counts as a digital measure of disease progression is ALS remains unexplored, despite successful use of similar endpoints in other neuromuscular diseases (23).

To address these gaps, we propose a simple and robust step counting algorithm that leverages ankle-worn accelerometer data. The algorithm builds on fundamental features of walking, i.e., periodicity, intensity, and cadence, and is designed to accommodate the heterogeneity, speed fluctuations, and asymmetries characteristic to ALS gait. Because our purpose was to develop an algorithm that quantifies gait in individuals that may use assistive devices, we relied solely on ankle-worn sensors. While wrist-worn accelerometers are commonly used in other populations (24–26), their application in ALS is limited due to interruptions in typical arm swings, that can lead to substantial errors when participants begin to use assistive devices.

We validate the method using several publicly available datasets of healthy adults as well as clinical and free-living data from individuals with ALS, capturing a range of walking speeds, assistive device use, and disease severity. Furthermore, we test the accuracy of the method to distinguish walking from other common daily activities (27). This multi-context validation framework enables analytical and clinical evaluation of algorithmic performance and supports the feasibility of ankle-worn step counting as a sensitive, interpretable, and scalable digital endpoint for mobility in ALS.

## 2. Methods

### 2.1. Population, devices, data

The proposed method was validated analytically and clinically using six independent datasets comprising of accelerometer data collected from healthy individuals and individuals diagnosed with ALS. Analytic validation was performed on five datasets, Human Physical Activity (*SPADES*) (28), Daily Life Activities (*DaLiAc*) (29), Identification of Walking, Stair Climbing, and Driving using Wearable Accelerometers (*IWSCD*) (30), Pedometer Evaluation Project (*PedEval*) (31), and Endurance Tasks to Measure Performance Fatigue in ALS (*PFALS*). Three of these datasets (*SPADES, DaLiAc,* and *IWSCD*) were used to study walking recognition, while two (*PedEval*, *PFALS*) were used to study step counting. The *SPADES*, *DaLiAc*, *IWSCD*, and *PedEval* datasets were identified in a public domain, while *PFALS* was collected by a research team at the Neurological Clinical Research Institute and Sean M. Healey & AMG Center for ALS at Massachusetts General Hospital (Boston, MA, USA). Clinical validation was performed on a dataset collected within the ALS Research Collaborative (*ARC*) led by the ALS Therapy Development Institute (Watertown, MA, USA). The *PFALS* and *ARC* datasets are not available in public domain due to privacy constraints, but data access can be requested by investigators (see **Data Availability**). The following sections describe available information on the investigated datasets; their summary is provided in **Table 1**.

**Table 1.** Demographics, body measures, and health status of participants involved in datasets included in this study. Age, height, weight, and BMI are provided as range (mean±SD), when available.

|  | *SPADES* | *DaLiAc* | *IUWCD* | *PedEval* | *PFALS* | *ARC* |
| --- | --- | --- | --- | --- | --- | --- |
| **Health status** | Healthy | Healthy | Healthy | Healthy | Healthy (15) / ALS (12) | ALS |
| **N** | 42 | 19 | 32 | 30 | 27 | 305 |
| **Sex (females)** | 15 | 8 | 19 | 15 | 8 / 2 | 103 |
| **Age (years)** | 18-30 (23.5±3.1) | 18-55 (26.5±7.7) | 23-52 (39.0±9.0) | 19-27 (21.9±5.4) | 44-68 (60.3±7.7) / 45-70 (59.9±7.4) | 20-78 (55.1±10.6) |
| **Height (cm)** | 151-180 (174.2±8.5) | 158-196 (177.0±11.1) | 147-193 (173.5±11.1) | 152-193 (171.0±10.8) | 158-183 (169.6±6.3) / 152-190 (175.3±11.9) | 152-203 (173.6±9.9) |
| **Weight (kg)** | 51-112 (73.8±15.0) | 54-108 (75.2±14.2) | 45-140 (77.0±22.9) | 43-136 (70.5±17.6) | 54-99 (78.2±13.2) / 57-103.9 (85.5±14.9) | 29.9-147 (79.1±15.8) |
| **BMI (kg/m2)** | 18-35 (24.7±4.1) | 17-34 (23.9±3.7) | 18-40 (25.2±5.6) | 17-37 (23.8±3.7) | 22-34 (27.7±4.3) / 22-40 (27.9±5.2) | 10-41 (26.2±4.5) |
| **Data collection** |  |  |  |  |  |  |
| **Environment** | Controlled | Controlled | Semi-controlled | Controlled & Semi-controlled | Controlled | Free-living |
| **Device** | ActiGraph GT9X | SHIMMER | ActiGraph GT3X+ | SHIMMER3 | APDM Opal | ActiGraph GT3X+ |
| **Body location** | Left ankle | Left ankle | Left ankle | Left ankle | Left foot | Left ankle |
| **Sampling rate** | 80 | 204.8 | 100 | 15 | 128 | 30 |
| **Disease characteristics** |  |  |  |  |  |  |
| **ALSFRS-R Q1—Q12** |  |  |  |  | 27-42 (34.9±4.1) | 12-48 (40.21±5.58) |
| **ALSFRS-R Q7** |  |  |  |  | 2-4 (3.2±0.7) | 0-4 (3.31±0.80) |
| **ALSFRS-R Q8** |  |  |  |  | 2-4 (2.5±0.67) | 1-4 (2.96±0.88) |
| **ALSFRS-R Q9** |  |  |  |  | 0-4 (1.5±1.4) | 0-4 (2.71±1.20) |

#### 2.1.1 Accuracy in walking recognition

##### SPADES

Forty-two participants (age: 23.5±3.1 years; 15 females) performed a variety of activities of daily living, including normal walking, ascending stairs, descending stairs, treadmill walk (1, 2, 3, and 3.5 mph), lying, sitting, standing, reclining, handwriting, computer typing, filling shelves, sweeping, running, cycling, and jumping jacks in supervised (laboratory) conditions. Raw accelerometer data were collected from participants’ ankles using ActiGraph GT9X (Pensacola, Florida) with sampling rate equal to 80 Hz and a dynamic range of ±8 g. Data were labelled by a research team member.

##### DaLiAc

Nineteen participants (age: 26.5±7.7 years; 8 females) performed thirteen activities including different postures (sitting, lying, standing), household activities (washing dishes, vacuuming, sweeping), walking behaviors (normal walking, running, stairs climbing), and sports activities (cycling, rope jumping). Raw accelerometer data were collected from participants’ ankles using the SHIMMER (Dublin, Ireland) with a sampling rate equal to 204.8 Hz and dynamic range of ±6 g. A dedicated smartphone app was used to label the beginning and ending time of each activity.

##### IUWCD

Twenty-four adults (age: 39.0±9.0 years; 19 females) participated in a study to identify walking, stair walking and driving from raw accelerometry data. The study included a walking trial (approximately 1 km) followed by a driving trial (approximately 21 km). The walking trial included walking on a level ground, up and down stairs, and up and down inclined paths. Immediately after the walking period, participants were accompanied to their vehicle, which they drove on a predefined route that included both highway and city driving. The walking trial lasted between 10 and 14 minutes while the driving trial lasted between 18 and 30 minutes, depending on traffic. Participants wore ActiGraph GT3X+ accelerometer on their left ankle that collected raw accelerometer data with sampling rate equal to 100Hz and a dynamic range of ± 6g.

#### 2.1.2 Accuracy in step counting

##### PedEval

Thirty participants (age: 21.9 ± 5.4 years, 15 females) performed three prescribed walking tasks: (1) a two-lap stroll along a designated path (continuous walk), (2) a scavenger hunt across four rooms (semi-continuous walk), and (3) a toy-assembling assignment using pieces distributed across a dozen bins located around a room (discontinuous walk). Participants were asked to wear the SHIMMER3 sensor on their left ankle. The sensor collected data with sampling frequency of 100 Hz and a dynamic range of ±4 g. Step counts were visually assessed and manually annotated by a research team member.

##### PFALS

The investigated dataset was collected as part of a pilot study of endurance tasks to measure performance fatigue in ALS. We analyzed data from 12 participants (age: 59.9±7.4 years, 2 females) with ALS and 15 healthy controls (age: 60.3±7.7 years, 8 females). In this task, participants were asked to wear the ADPM Opal devices (Estenfeld, Germany) at their ankles, and walk at about 75% of their maximum walking speed back and forth for up to 20 minutes along the 10-meter course, completing 180 degree turns at the cones positioned at each end. Each trip between the cones counted as one lap. In this study, data from the initial six laps of the trial was evaluated for all participants. Devices recorded continuous triaxial accelerometer measures as a sampling frequency equal to 128 Hz and a dynamic range of ±2 g. This study was approved by the Institutional Review Board of Massachusetts General Brigham (IRB# 2020P000546).

#### 2.1.3. Accuracy in distinguishing disease status

##### ARC

We analyzed data from 305 participants (age: 55.1±10.6years, 103 females) diagnosed with ALS wearing ActiGraph GT3X+ accelerometers on their left ankle. Devices recorded continuous triaxial accelerometer measurements at a sampling frequency of 30 Hz with a dynamic range of ±6 g. In this free-living observation, participants were instructed to wear the devices as much as possible for 7 days excluding bathing.

This study adhered to the guidelines outlined in the Declaration of Helsinki – Ethical Principles for Medical Research Involving Human Subjects. The study protocol was approved by the Institutional Review Board (ADVARRA Center for IRB Intelligence). All participants provided informed consent prior to any study procedures. There was no compensation for participation in the study.

### 2.2. Algorithm description

The proposed step counting method leverages the observation that walking consists of repetitive, oscillatory movements characterized by distinct periodicity and amplitude, particularly when captured by worn accelerometers. These periodic features arise from biomechanical events such as heel strikes, which produce transient, high-amplitude changes in acceleration along the axis aligned with the shin (**Figure 1A**). To discriminate walking from other daily activities and to detect individual steps in heterogeneous free-living conditions, we apply a continuous wavelet transform (CWT) to the raw accelerometer signal. This transform decomposes the signal into a time-frequency representation, allowing for the detection of localized periodic events while being robust to signal non-stationarity (**Figure 1B**). By focusing on the signal’s vertical component, i.e., where heel strikes are reflected as sharp spikes, our method provides a robust framework for detecting gait in individuals with varying movement patterns and gait abnormalities.

**Figure 1.**
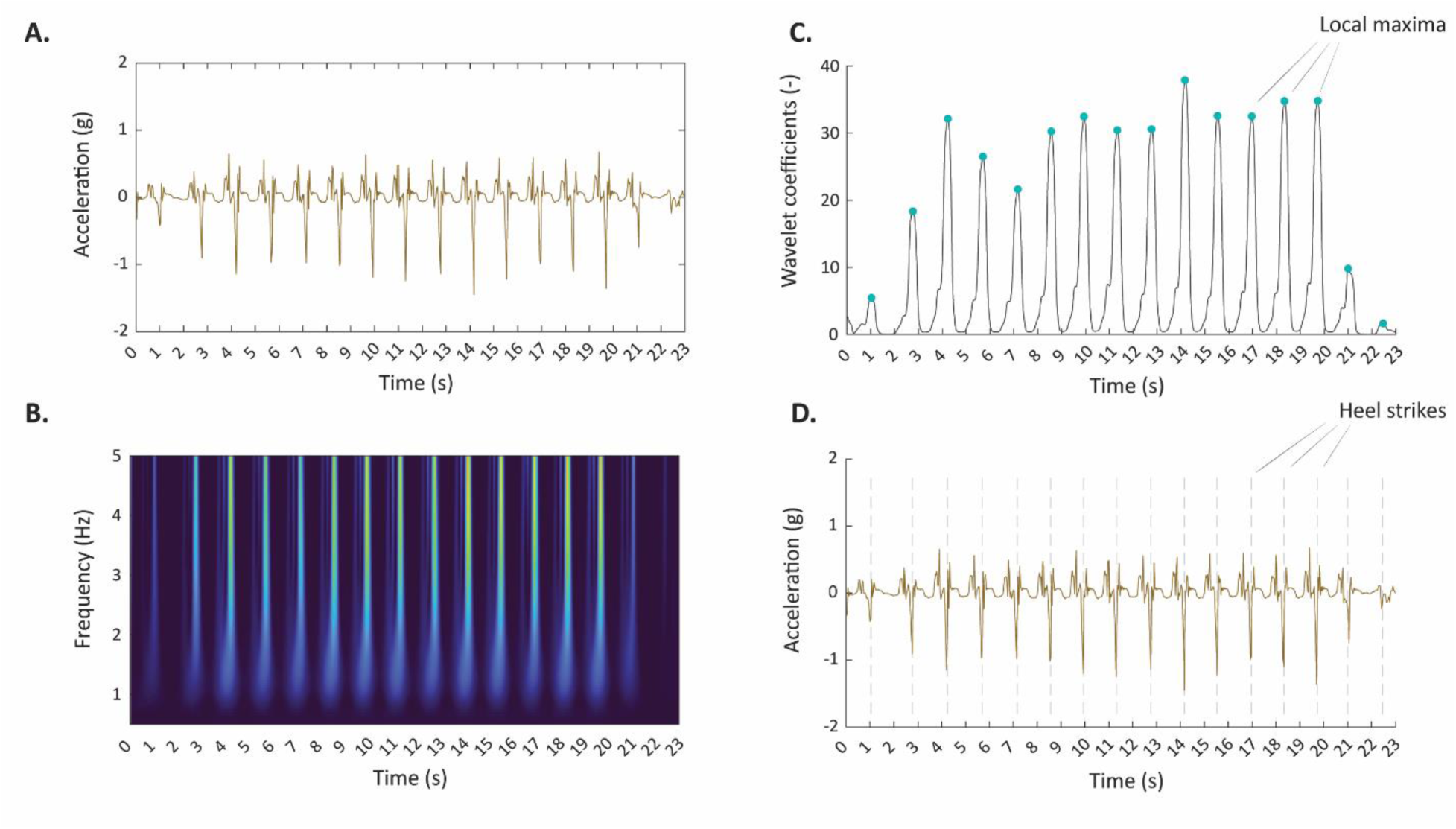
Framework for detecting heel strikes using ankle-worn accelerometers. Signal from vertical axis (**A**) is decomposed using a continuous wavelet transform which uncovers sharp spikes corresponding to individual heel strikes (**B**). Local peaks in frequency-wise summation of wavelet coefficients help identify heel strikes locations within the signal s(**C**). Identified locations correspond to heel strikes from one foot (**D**); assuming another foot making the same number of steps, total step counts are calculated as number of heel strikes multiplied by 2.

In our method, the CWT is implemented using the generalized Morse wavelet, defined as 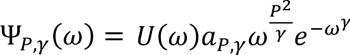 parameterized with a symmetry constant γ = 3, ensuring zero skewness, and a time-bandwidth product P² = 10, which balances time and frequency localization (**Supplementary** Figure 1). This configuration allows the wavelet coefficients to be particularly sensitive to abrupt changes in signal amplitude, such as those produced during heel strikes.

In our approach, wavelet coefficients (product of CWT) are computed across a physiologically relevant frequency range (up to 5Hz), and their values are summed at each time point to enhance the signal-to-noise ratio of periodic gait-related events (**Figure 1C**). We identify local peaks in the resulting time series of summed coefficients and classify them as heel strikes if they exceed a fixed threshold set at 0.1 (**Figure 1D**).

To prevent spurious peaks (e.g., from isolated stomps or noise) from being misclassified as steps, we apply additional temporal time constraints. Specifically, our algorithm enforces that timestamps *t*_*i*_, *t*_*i*+1_, …, *t*_*i*+*k*_ of consecutive heel strikes from the same leg must be separated by time intervals Δ_*i*_= *t*_*i*+1_ − *t*_*i*_ with physiologically plausible Δ_*min*_= 0.85 seconds and Δ_*max*_= 2.5 seconds, corresponding to a very fast (about 2.35 steps per second) and a very slow walking speed (0.8 steps per second), respectively. Furthermore, the difference between consecutive between-strike intervals δ_*i*_ = |Δ_*i*+1_ − Δ_*i*_| should not exceed 0.5 seconds to further exclude implausible or artifact-driven detections.

In the analytical validation, we focus on two gait-related measures: walking activity time and step counts. Walking activity is identified for all nonsingular heel strikes if ∀_*j*_∈ {*i*, …, *i* + *k* − 1}, *t*_*i*+1_ − *t*_*i*_ ≤ 3 seconds. Steps were included in the step count outcome measure if they were classified as walking activity. Assuming another foot making the same number of steps, total step counts were computed as number of detected heel strikes multiplied by 2. Walking activity recognition rate was investigated to determine potential activities of daily living that might inflate step counts in free-living observations.

### 2.4. Statistical analysis

All raw accelerometer data was resampled to 10 Hz using linear interpolation prior to analysis to ensure consistent temporal resolution across datasets (32). This sampling rate was previously demonstrated as sufficient to capture gait characteristics across various devices and sensor body locations (27). Walking periods were identified in physical activity data from three annotated datasets (*SPADES*, *DaLiAc*, and *IUWCD*). To evaluate analytical validity of our method for walking recognition, we compared the algorithm-derived walking segments to reference activity labels provided in the source datasets. Sensitivity and specificity metrics were calculated for each trial, averaged across repeated measures within subject, and then aggregated across participants. Sensitivity was defined as the proportion of correctly identified walking segments among all annotated walking episodes (including level walking, stair climbing, and treadmill walking), while specificity was calculated as the proportion of correctly excluded non-walking segments (e.g., sitting, standing, household tasks). We report sensitivity as mean and 95 % confidence intervals (CI).

We assessed the accuracy of step counting using Bland-Altman analyses across two validation settings. Depending on the dataset, ground truth was assessed visually and manually annotated by research team members either by following a walking person (*PedEval*) or by evaluating video recording (*PFALS*). In the latter, step counts were assessed as a mean score assessed by two co-authors of this paper (KMB and KTC). We report the accuracy for step counting in terms of mean error (bias) and levels of agreement (LoA).

In the clinical validation phase, we utilized data collected over a one-week monitoring period (*ARC*). Step counts were aggregated on the daily level for days with at least 16 hours of wear time, as determined by non-wear detection algorithm proposed by Choi and colleagues (33), and averaged if there were at least 3 days of days with sufficient sensor wear-time.

Two sample *t*-tests were used to compare step counts with ALSFRS-R gross motor function questions regarding turning in bed and adjusting bed clothes (Q7), walking (Q8), and climbing stairs (Q9). Each test considered step counts observed in participants reporting normal functioning and those reporting decreased functioning. Statistics on demographics, health descriptions, and performed activities are reported using mean and standard deviations (SD).

## 3. Results

The aggregated dataset used to validate the method comprised of data from 452 individuals, including 138 healthy individuals and 314 individuals diagnosed with ALS. Data from 93 individuals were used to determine the accuracy to recognize walking activity, data from 57 (45 healthy, 12 ALS) individuals were used for analytical validation of step counting, while data from 302 individuals were used to clinical validation in ALS.

Altogether we analyzed data from 80,989 hours of activity, including over 21 hours for analytical validation of walking recognition (8.75 h of walking activities, 12.65 h of non-walking activities), 15.57 hours for analytical validation of step counting, and over 80,952 hours (about 3373 days) for clinical validation in ALS.

### 3.1. Analytical validation

#### 3.1.1 Walking recognition accuracy

Walking recognition was highly accurate for a wide range of walking activities, such as normal walking (e.g., in *SPADES* the mean sensitivity was 0.995, CI = [0.997, 0.999]), ascending stairs (0.947 [0.908, 0.987]), descending stairs (0.915 [0.847, 0.983]), or walking on a treadmill at 1 mph (0.983 [0.978, 0.987]) (Figure 2A). Our method also accurately recognized non-walking activities, including repetitive activities, such as cycling (e.g., in *DaLiAc* mean specificity was equal 1 [1, 1]) or running (0.981 [0.951, 1]) (Figure 2B). Our results showed that car driving data collected in *IUWCD*, often represented as high-frequency vibrations (34), was entirely classified as non-walking (1 [1, 1]) (Figure 2C). Decreased specificity was observed for sweeping the floor (in *DaLiAc* 0.851 [0.815, 0.887]) and vacuuming (0.877 [0.814, 0.940]). However, it is unknown to the authors whether these activities in fact consisted of entirely non-walking movements. Finally, systematically decreased specificity was reported for jumping (in *SPADES* 0.669 [0.545, 0.792], in *DaLiAc* 0.852 [0.770, 0.934]).

**Figure 2.**
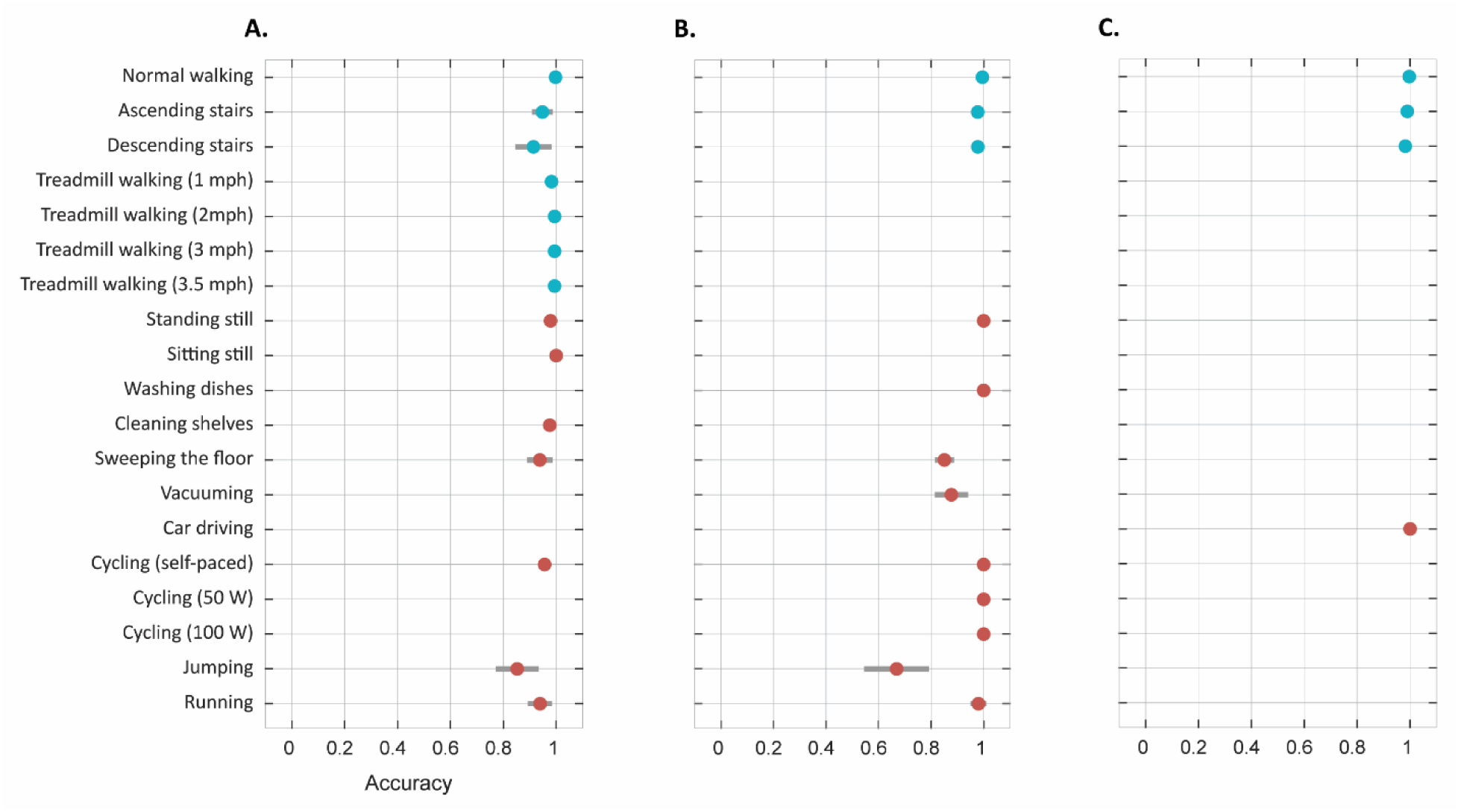
Sensitivity (blue) and specificity (red) estimated for walking recognition in populations of ambulatory participants investigated in *SPADES* (A), *DaLiAc* (B), and *IUWCD* (C) studies.

#### 3.1.2. Step counting accuracy

Altogether, according to the ground truth, participants included in *PedEval* performed a mean (SD) of 1045.73 (85.81) steps during continuous, 681.91 (88.37) steps during semi-continuous, and 180.81 (16.65) steps during discontinuous walking activities, respectively. The corresponding estimations calculated using our method were 1048.15 (84.89) steps for continuous, 660.67 (93.45) steps for semi-continuous, and 190.29 (18.29) steps for discontinuous walking activities.

Using Bland-Altman analysis, we determined that the mean bias (LoA) was equal to 0.42 (−11.60, 12.44) or 0.04 % for continuous walking (Figure 3A), −4.33 (−61.81, 53.15) or 0.64 % for semi-continuous walking (Figure 3B), and 1.36 (−44.45, 47.17) or 0.76 % for discontinuous walking (Figure 3C), indicating excellent reliability of the algorithm on a population level. The mean (SD) ALSFRS-R total score of individuals participating in *PFALS* dataset was equal to 34.9 (4.1), mean response to Q7 was equal to 3.2 (0.7), Q8 2.5 (0.7), and Q9 1.5 (1.4).

**Figure 3.**
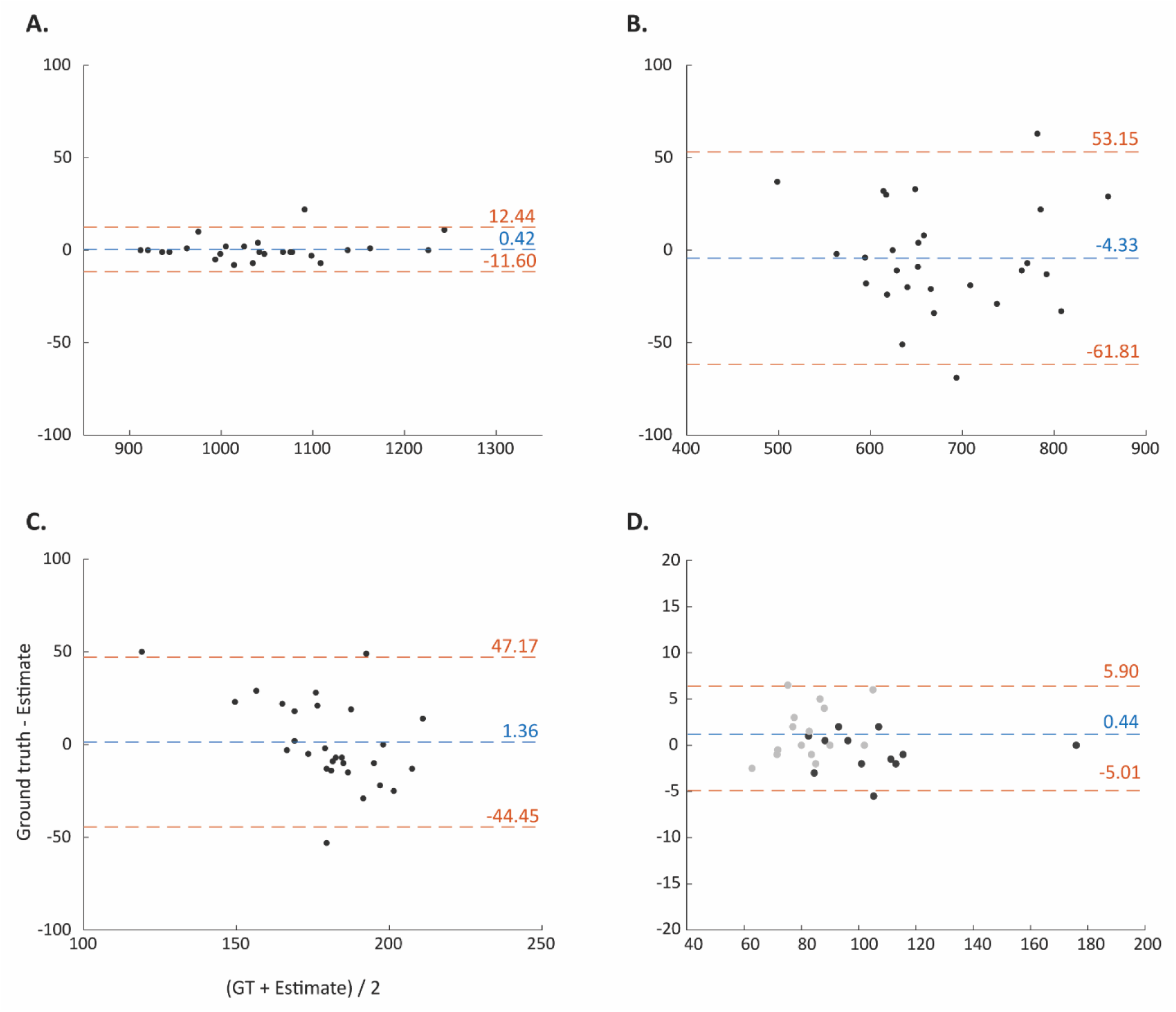
Bland-Altman plots on step counting accuracy in three walking scenarios investigated in *PedEval* dataset (continuous walk A, semi-continuous walk B, and discontinuous walk C) and one walking scenario investigated in *PFALS* dataset (D). In each scenario, ground truth (GT) was assessed by a visual observer. Black dots correspond to observations from healthy individuals, grey dots correspond to observations from individuals diagnosed with ALS.

In *PFALS*, ALS participants and healthy controls performed a similar number of steps, i.e., 1269 and 1249, respectively. In this controlled experiment, our method accurately counted steps regardless of participants’ health conditions. In healthy participants, mean bias (LoA) was equal to −0.75 (−5.06, 3.56) or −1.75 %, while in ALS participants mean bias was equal to 1.40 (−4.28, 7.08) or 0.4 %. Cumulatively, mean bias (LoA) was equal to 0.44 (54.01, 5.90) or 0.56 %, suggesting accurate step counting performance across the two populations (Figure 3D).

### 3.2. Clinical validation

Clinical validation data were collected between September 2014 and January 2023 on 305 participants diagnosed with ALS. There were 251 participants who met the data inclusions criteria (at least 3 days with a minimum wear-time of 16 hours). The mean (SD) data collection period was equal to 13.44 (8.44) days, while step counts were estimated for a mean (SD) of 5.13 (1.89) days. At baseline, study participants reported a mean (SD) ALSFRS-R total score equal to 40.21 (5.58), while their mean (SD) responses to Q7, Q8, and Q9 were equal to 3.31 (0.80), 2.96 (0.88), and 2.76 (1.20), respectively.

Using step counts, we observed statistically significant differences between participants reporting normal gross motor functioning and those reporting impairments in this domain (Figure 4). For example, participants reporting “normal” gait (Q8) performed a mean of 3984 [3537, 4432] step counts per day, significantly more compared to those reporting “early ambulation difficulties” (3197 [2837, 3557], *p* = 0.009), and “walking with assistance” (1283 [1063, 1503], *p* = <0.001). Importantly, the algorithm estimated almost no steps for those reporting “nonambulatory functional movement” (81 [12, 150], *p* = <0.001), indicating that it is robust to non-walking activities of daily living.

**Figure 4.**
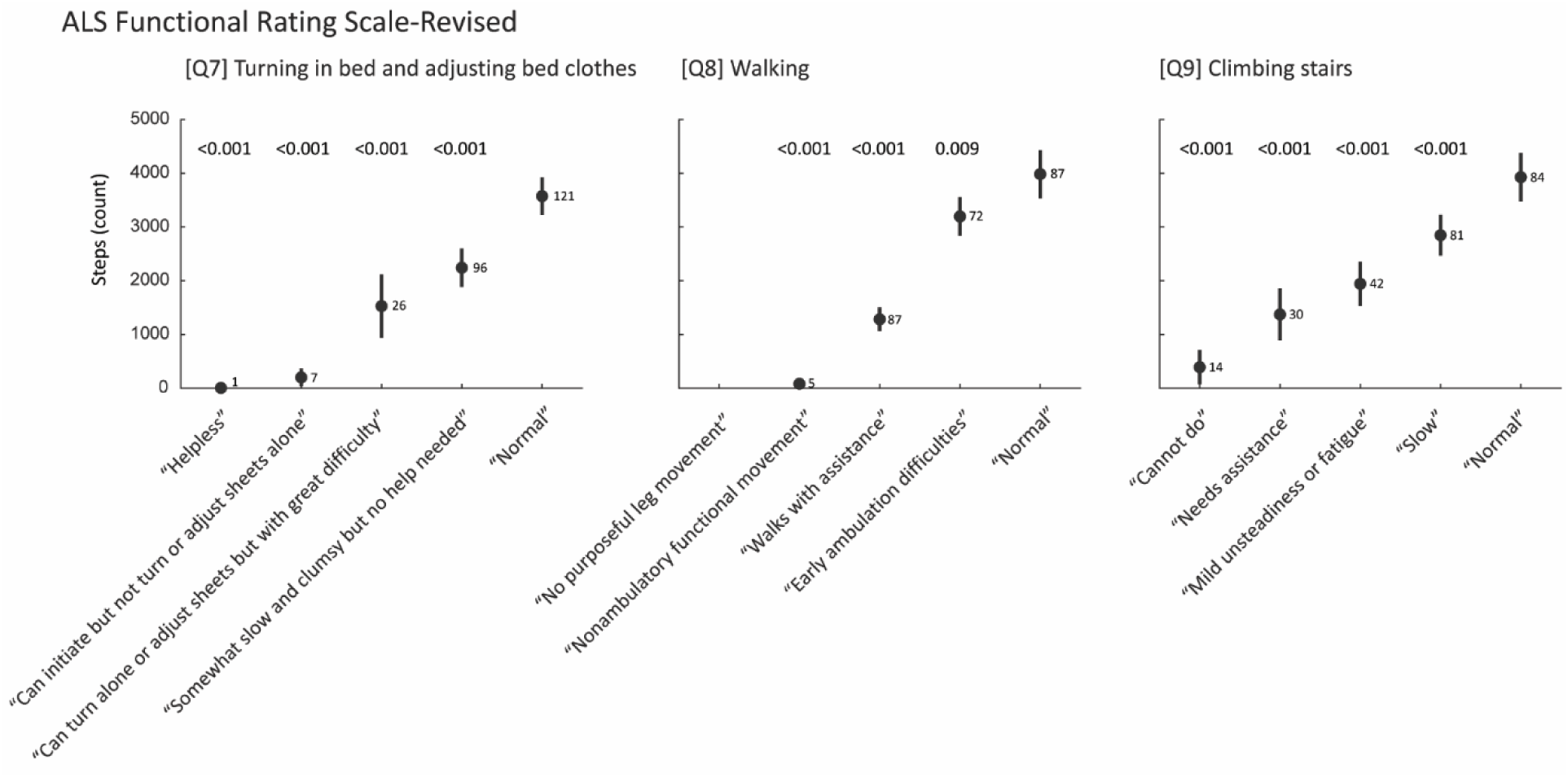
Mean and 95% CI of step counts stratified by responses to ALSFRS-R questions on gross motor functioning subscale (Q7—Q9) along with *p*-values of two sample t-test (i.e., step counts observed in participants reporting normal functioning and those reporting decreased functioning). Values displayed next to the mean step counts indicate group size reporting a given functional status.

## 4. Discussion

This study demonstrates the validity and applicability of a step-counting method for quantifying ambulatory activity in individuals with ALS. By leveraging a continuous wavelet transform to detect periodic peaks corresponding to heel strikes, our algorithm offers a robust and interpretable approach for step detection using accelerometers placed on a foot or ankle, body location which appears favorable to capture gait events in individuals using assistive devices (**Supplementary** Figure 2 **panels B—D**). We determined the utility of the method following principles of the V3 framework, which underscores the need for analytical and clinical validation of sensor-based digital health technologies in populations with changing health status (35,36).

Our analytical validation was conducted in two ways, (1) focusing on the accuracy of the method to identify periods of walking activity and (2) counting steps within these periods. The walking recognition accuracy was verified using three independent, publicly available datasets. It was determined to be highly sensitive to walking activities and highly specific to most non-walking activities defined by the respective dataset authors. Notably, a marginally decreased specificity was observed for selected household activities, such as vacuuming or sweeping the floor. These activities, however, might include a small number of steps and a certain level of misclassification is warranted. Importantly, the method remained insensitive to activities that might present in the data as repetitive spikes, such as cycling or driving a car.

The proposed approach also demonstrated accurate estimation of step counts both in controlled and semi-controlled assessments. In each investigated scenario the mean bias was close to zero, while wider levels of agreement were calculated for semi-controlled tasks. This variation may be attributed to complex structure of movement patterns, such as hesitations, pauses, or weight-bearing steps that can produce signal artifacts resembling step-like events. Additionally, the absence of a standardized protocol for specific transitions, such as turning around, navigating obstacles, or climbing stairs, may have introduced variability in recorded signals that contributed to reported misclassification. These factors highlight inherent challenges of step counting in naturalistic settings where movements often deviate from steady, linear gait patterns.

For clinical validation, we evaluated whether the method provides clinically meaningful insights into mobility patterns in people living with ALS, a population characterized by progressive motor decline and high interindividual variability in gait. Our results show that the method can differentiate step counts between individuals with normal gait, early-stage ambulatory difficulties, and those requiring assistance. This distinction aligns with known clinical trajectories in ALS and demonstrates the method’s ability to detect functionally and clinically relevant changes in walking behavior, which is a critical feature for a potential digital outcome measure used in a therapeutic evaluation or clinical trial. According to our knowledge, this is the first study to assess step count levels in a large-scale free-living observation involving individuals with ALS. In our previous study (5), we enrolled twenty people living with ALS whose baseline daily step counts were estimated as 1871 [1137, 2605] steps, substantially lower than the step counts reported in this study. The lower step counts in the earlier cohort may reflect participants in a more advanced disease stage, as indicated by a substantially lower average ALSFRS-R total score (mean 31.40) compared to the group reported here (mean 40.21).

Our findings align with prior work which emphasizes that ankle-worn devices are particularly suited for step detection in populations with altered gait patterns, including those with neurodegenerative diseases and requiring assistance. It is worth highlighting that our method detects heel strikes using data collected with a low sampling frequency (10 Hz), in contrast to many commercial algorithms that require higher sampling frequencies to achieve comparable accuracy (22,37). This property is especially important in free-living observations of individuals with ALS, where slower gait speeds, the use of assistive devices (21), or variable walking surfaces (27) may introduce artefacts that hinder approaches relying on time-domain signal analyses, and where employing more sophisticated devices may not be feasible. The clear distinction in step counts observed between participants with early ambulatory impairment and those requiring assistance (3197 [2837, 3557] vs. 1283 [1063, 1503]) underscores the method’s sensitivity to clinically relevant differences in mobility status, reinforcing its utility as a candidate digital endpoint for ALS studies.

Several limitations should also be acknowledged. First, the method assumes a basic level of periodicity and intensity in the accelerometer signal, which may lead to errors in contexts involving sporadic or irregular stepping, such as turning, standing, or shuffling. This challenge is compounded by the lack of consensus on how to define “walking” in real-world settings, as it remains unclear whether a single step, two steps, or a continuous sequence of steps should be classified as walking (27). Second, due to the secondary nature of this analysis and the absence of a standardized turning protocol in the original datasets (e.g., in *PFALS*), we were unable to assess the algorithm’s performance specifically during transitions such as straight walks versus turning. Third, while the algorithm provided meaningful indications across the included ALS datasets, the free-living observations did not include ground truth step counts, limiting our ability to assess its accuracy under naturalistic conditions. Future research should explore these limitations but also consider the integration of additional sensor modalities (e.g., gyroscopes, magnetometers) to capture a broader range of gait-related metrics, including stride length, cadence variability, and measures of balance.

## 5. Conclusions

The reliance of our method on the fundamental properties of the gait signal, recorded from the ankle, rather than the wrist, supports its applicability across diverse measurement conditions. By validating the algorithm across multiple independent datasets, including those collected in controlled, semi-controlled, and free-living settings, we demonstrated its reproducibility and potential for use in real-world studies. Importantly, the algorithm accurately counts steps and walking activity in healthy participants and those with ALS who have gait disturbances – even those requiring assistance. This overcomes a major existing shortcoming of gait quantification in ALS trials – the floor effect driven by the inability to accurately count steps as gait deteriorates. With this method, researchers can more assuredly employ accelerometry in clinical trials. We have made our algorithm open-source to enhance transparency and support adoption by researchers in ALS and other populations. The method can also be adapted for other devices, or research questions, as needed. Together, these attributes position the method as a promising foundation for future development and application of digital outcome measures on mobility in ALS and other diseases characterized by deterioration of gait.

## 6. List of abbreviations

ALSFRS-R: ALS Functional Rating Scale-Revised.
ARC: ALS Research Collaborative dataset.
BMI: Body mass index.
CI: confidence intervals.
CWT: continuous wavelet transform.
DaLiAc: Daily Life Activities dataset.
DHT: Digital Health Technology.
GT: ground truth.
IWSCD: Identification of Walking, Stair Climbing, and Driving using Wearable Accelerometers dataset.
LoA: levels of agreement.
PedEval: Pedometer Evaluation Project dataset.
PFALS: Endurance Tasks to Measure Performance Fatigue in ALS dataset.
SD: standard deviation.
SPADES: Human Physical Activity dataset.

## 7. Declarations

### 7.1 Ethics approval and consent to participate

This study adhered to the guidelines outlined in the Declaration of Helsinki – Ethical Principles for Medical Research Involving Human Subjects. The study protocol for *ARC* was approved by the Institutional Review Board (ADVARRA Center for IRB Intelligence). The study protocol for *PFALS* was approved by the Institutional Review Board of Massachusetts General Brigham (IRB# 2020P000546). In *ARC* and *PFALS* all participants provided informed consent prior to any study procedures.

### 7.2 Consent for publication

Not applicable.

### 7.3 Availability of data and materials

#### 7.3.1 Code availability

Methods used in this study are publicly available under the following link: **(link to be provided upon manuscript acceptance).**

### 7.6 Authors contribution

MS proposed the method, identified open-source datasets, processed the data, performed statistical analysis, created figures, prepared the first draft.

KMB designed the *PFALS* project, collected data in the *PFALS* project, provided clinical oversight, reviewed the analyses and manuscript drafts.

KTC collected data in the PFALS project, provided clinical oversight, reviewed the analyses and manuscript drafts. NC designed the *PFALS* project, provided clinical oversight, reviewed the analyses and manuscript drafts.

SM provided clinical oversight, reviewed the analysis and manuscript drafts. AP designed the *ARC* project, reviewed the analyses and manuscript drafts. FV designed the *ARC* project, reviewed the analyses and manuscript drafts.

JDB designed the *PFALS* project, provided clinical oversight, reviewed the analyses and manuscript drafts.

## Data Availability

The PFALS and ARC datasets may be shared upon request and after review and approval by the owners of the data. Shared data will consider deidentified summary statistics used for analytical analysis. Related correspondence should be sent to (PFALS) and (ARC).

## Acknowledgements

The authors are grateful to participants and their caregivers. They also appreciate researchers who made their datasets publicly available.

## References

1. Fritz S, Lusardi M. White paper: “walking speed: the sixth vital sign”. J Geriatr Phys Ther. 2009;32(2):46–9.

2. Studenski S, Perera S, Patel K, Rosano C, Faulkner K, Inzitari M, et al. Gait Speed and Survival in Older Adults. JAMA [Internet]. 2011;305(1):50–8. Available from: 10.1001/jama.2010.1923

3. Middleton A, Fritz SL, Lusardi M. Walking speed: the functional vital sign. J Aging Phys Act. 2015 Apr;23(2):314–22.

4. Radovanović S, Milićev M, Perić S, Basta I, Kostić V, Stević Z. Gait in amyotrophic lateral sclerosis: Is gait pattern differently affected in spinal and bulbar onset of the disease during dual task walking? Amyotroph Lateral Scler Frontotemporal Degener. 2014 Dec;15(7–8):488–93.

5. Johnson SA, Karas M, Burke KM, Straczkiewicz M, Scheier ZA, Clark AP, et al. Wearable device and smartphone data quantify ALS progression and may provide novel outcome measures. npj Digit Med [Internet]. 2023;6(1):34. Available from: 10.1038/s41746-023-00778-y

6. Karas M, Olsen J, Straczkiewicz M, Johnson SA, Burke KM, Iwasaki S, et al. Tracking amyotrophic lateral sclerosis disease progression using passively collected smartphone sensor data. Ann Clin Transl Neurol [Internet]. 2024;11:1380–92. Available from: https://onlinelibrary.wiley.com/doi/abs/10.1002/acn3.52050

7. Colón-Emeric CS, McDermott CL, Lee DS, Berry SD. Risk Assessment and Prevention of Falls in Older Community-Dwelling Adults: A Review. JAMA. 2024 Apr;331(16):1397–406.

8. Verghese J, Holtzer R, Lipton RB, Wang C. Quantitative Gait Markers and Incident Fall Risk in Older Adults. Journals Gerontol Ser A [Internet]. 2009;64A(8):896–901. Available from: 10.1093/gerona/glp033

9. Schniepp R, Huppert A, Decker J, Schenkel F, Schlick C, Rasoul A, et al. Fall prediction in neurological gait disorders: differential contributions from clinical assessment, gait analysis, and daily-life mobility monitoring. J Neurol [Internet]. 2021;268(9):3421–34. Available from: 10.1007/s00415-021-10504-x

10. Beswick E, Fawcett T, Hassan Z, Forbes D, Dakin R, Newton J, et al. A systematic review of digital technology to evaluate motor function and disease progression in motor neuron disease. J Neurol. 2022 Dec;269(12):6254–68.

11. Chipika RH, Finegan E, Li Hi Shing S, Hardiman O, Bede P. Tracking a Fast-Moving Disease: Longitudinal Markers, Monitoring, and Clinical Trial Endpoints in ALS. Front Neurol [Internet]. 2019;Volume 10. Available from: https://www.frontiersin.org/journals/neurology/articles/10.3389/fneur.2019.00229

12. Proudfoot M, Jones A, Talbot K, Al-Chalabi A, Turner MR. The ALSFRS as an outcome measure in therapeutic trials and its relationship to symptom onset. Amyotroph Lateral Scler Front Degener [Internet]. 2016;17(5–6):414–25. Available from: 10.3109/21678421.2016.1140786

13. Cedarbaum JM, Stambler N, Malta E, Fuller C, Hilt D, Thurmond B, et al. The ALSFRS-R: a revised ALS functional rating scale that incorporates assessments of respiratory function. BDNF ALS Study Group (Phase III). J Neurol Sci. 1999 Oct;169(1–2):13–21.

14. Blair HA. Tofersen: First Approval. Drugs. 2023 Jul;83(11):1039–43.

15. Smith SE, McCoy-Gross K, Malcolm A, Oranski J, Markway JW, Miller TM, et al. Tofersen treatment leads to sustained stabilization of disease in SOD1 ALS in a “real-world” setting. Ann Clin Transl Neurol. 2025 Feb;12(2):311–9.

16. Ruffo P, Traynor BJ, Conforti FL. Advancements in genetic research and RNA therapy strategies for amyotrophic lateral sclerosis (ALS): current progress and future prospects. J Neurol. 2025 Feb;272(3):233.

17. Straczkiewicz M, Burke KM, Calcagno N, Premasiri A, Vieira FG, Onnela JP, et al. Free-living monitoring of ALS progression in upper limbs using wearable accelerometers. J Neuroeng Rehabil [Internet]. 2024;21(1):223. Available from: 10.1186/s12984-024-01514-7

18. Gupta AS, Patel S, Premasiri A, Vieira F. At-home wearables and machine learning sensitively capture disease progression in amyotrophic lateral sclerosis. Nat Commun [Internet]. 2023;14(1):5080. Available from: 10.1038/s41467-023-40917-3

19. Vieira FG, Venugopalan S, Premasiri AS, McNally M, Jansen A, McCloskey K, et al. A machine-learning based objective measure for ALS disease severity. npj Digit Med [Internet]. 2022;5(1):45. Available from: 10.1038/s41746-022-00588-8

20. Musson LS, Mitic N, Leigh-Valero V, Onambele-Pearson G, Knox L, Steyn FJ, et al. The Use of Digital Devices to Monitor Physical Behavior in Motor Neuron Disease: Systematic Review. J Med Internet Res [Internet]. 2025 Apr;27:e68479. Available from: 10.2196/68479

21. Tsamassiotis S, Schwarze M, Gehring P, Karkosch RF, Tücking LR, Einfeldt AK, et al. Accelerometers can correctly count orthopaedic patients’ early post-operative steps while using walking aids. J Exp Orthop [Internet]. 2025;12(1):e70134. Available from: https://esskajournals.onlinelibrary.wiley.com/doi/abs/10.1002/jeo2.70134

22. Pan J, Wei S. Accuracy and reliability of accelerometer-based pedometers in step counts during walking, running, and stair climbing in different locations of attachment. Sci Rep [Internet]. 2024;14(1):27761. Available from: 10.1038/s41598-024-78684-w

23. Servais L, Eggenspieler D, Poleur M, Grelet M, Muntoni F, Strijbos P, et al. First regulatory qualification of a digital primary endpoint to measure treatment efficacy in DMD. Vol. 29, Nature medicine. United States; 2023. p. 2391–2.

24. Koffman L, Crainiceanu C, Muschelli J. Comparing Step Counting Algorithms for High-Resolution Wrist Accelerometry Data in NHANES 2011-2014. Med Sci Sports Exerc. 2024;

25. Small SR, Chan S, Walmsley R, VON Fritsch L, Acquah A, Mertes G, et al. Self-Supervised Machine Learning to Characterize Step Counts from Wrist-Worn Accelerometers in the UK Biobank. Med Sci Sports Exerc. 2024 Oct;56(10):1945–53.

26. Del Pozo Cruz B, Ahmadi M, Naismith SL, Stamatakis E. Association of Daily Step Count and Intensity With Incident Dementia in 78 430 Adults Living in the UK. JAMA Neurol. 2022 Oct;79(10):1059–63.

27. Straczkiewicz M, Huang EJ, Onnela JP. A “one-size-fits-most” walking recognition method for smartphones, smartwatches, and wearable accelerometers. npj Digit Med [Internet]. 2023;6(1):29. Available from: 10.1038/s41746-022-00745-z

28. John D, Tang Q, Albinali F, Intille S. An Open-Source Monitor-Independent Movement Summary for Accelerometer Data Processing. J Meas Phys Behav [Internet]. 2(4):268–81. Available from: http://journals.humankinetics.com/view/journals/jmpb/2/4/article-p268.xml

29. Leutheuser H, Schuldhaus D, Eskofier BM. Hierarchical, Multi-Sensor Based Classification of Daily Life Activities: Comparison with State-of-the-Art Algorithms Using a Benchmark Dataset. PLoS One [Internet]. 2013;8(10):1–11. Available from: 10.1371/journal.pone.0075196

30. Karas M, Straczkiewicz M, Fadel W, Harezlak J, Crainiceanu CM, Urbanek JK. Adaptive empirical pattern transformation (ADEPT) with application to walking stride segmentation. Biostatistics [Internet]. 2019;22(2):331–47. Available from: 10.1093/biostatistics/kxz033

31. Mattfeld R, Jesch E, Hoover A. A new dataset for evaluating pedometer performance. In: 2017 IEEE International Conference on Bioinformatics and Biomedicine (BIBM). 2017. p. 865–9.

32. Straczkiewicz M, Keating NL, Thompson E, Matulonis UA, Campos SM, Wright AA, et al. Open-Source, Step-Counting Algorithm for Smartphone Data Collected in Clinical and Nonclinical Settings: Algorithm Development and Validation Study. JMIR Cancer [Internet]. 2023 Nov;9:e47646. Available from: http://www.ncbi.nlm.nih.gov/pubmed/37966891

33. Choi L, Liu Z, Matthews CE, Buchowski MS. Validation of accelerometer wear and nonwear time classification algorithm. Med Sci Sports Exerc. 2011 Feb;43(2):357–64.

34. Straczkiewicz M, Urbanek JK, Fadel WF, Crainiceanu CM, Harezlak J. Automatic car driving detection using raw accelerometry data. Physiol Meas. 2016;37(10).

35. Goldsack JC, Coravos A, Bakker JP, Bent B, Dowling A V, Fitzer-Attas C, et al. Verification, analytical validation, and clinical validation (V3): the foundation of determining fit-for-purpose for Biometric Monitoring Technologies (BioMeTs). npj Digit Med [Internet]. 2020;3(1):55. Available from: 10.1038/s41746-020-0260-4

36. Bakker JP, Barge R, Centra J, Cobb B, Cota C, Guo CC, et al. V3+ extends the V3 framework to ensure user-centricity and scalability of sensor-based digital health technologies. npj Digit Med [Internet]. 2025;8(1):51. Available from: 10.1038/s41746-024-01322-2

37. McCullagh R, Dillon C, O’Connell AM, Horgan NF, Timmons S. Step-Count Accuracy of 3 Motion Sensors for Older and Frail Medical Inpatients. Arch Phys Med Rehabil [Internet]. 2017 Feb 1;98(2):295–302. Available from: 10.1016/j.apmr.2016.08.476

