## Supplementary Materials for "Analytical and clinical validation of step counting method in people living with amyotrophic lateral sclerosis"

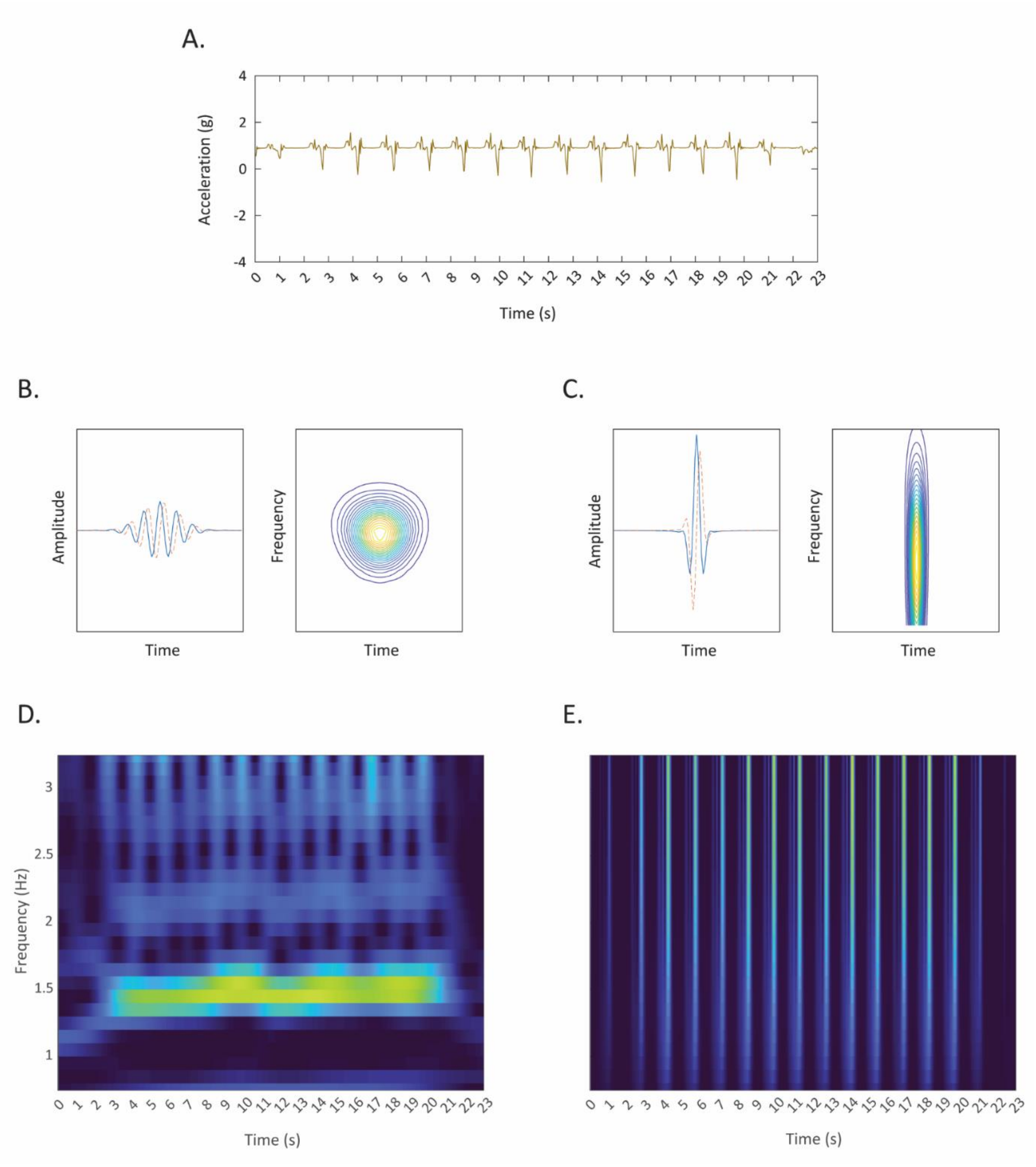


**Supplementary Figure 1.** Selection of mother wavelet impacts identification of heel strikes. Raw accelerometer data collected in vertical axis using ankle-worn device (A) may be decomposed using Morse wavelet with symmetry parameter $\gamma = 3$ and a time-bandwidth product $P^2 = 60$ (B) or $\gamma = 3$ and $P^2 = 10$ (C). The narrower frequency response may be used to capture step frequency, e.g., in regular flat walking (D) while wider frequency response may be used to capture heel strikes (E), as presented in our method.

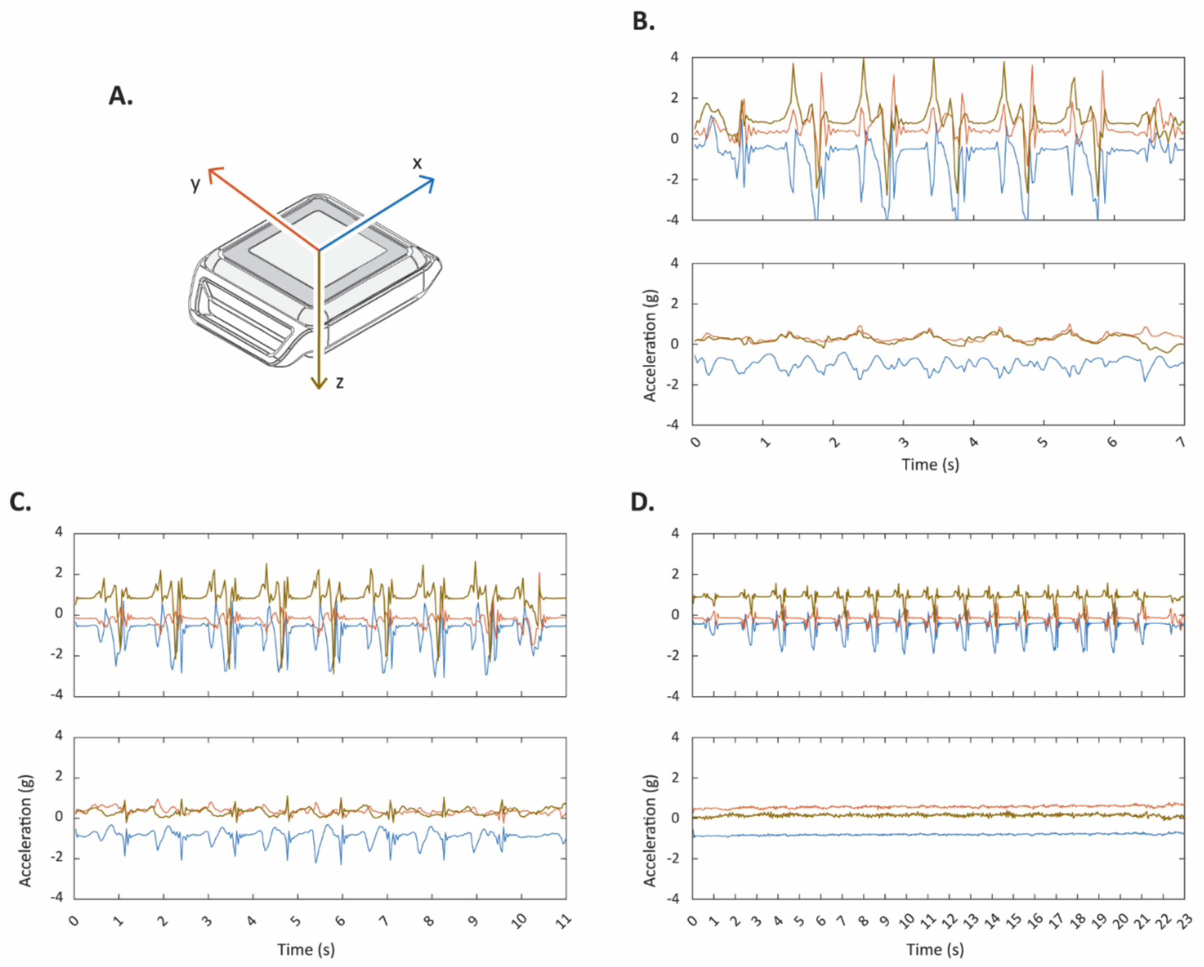


**Supplementary Figure 2.** Coordinate system of the measurement device (A) along with raw accelerometer data collected by foot- (top panel) and wrist-worn (bottom panel) devices for three participants observed in the *PFALS* dataset: healthy control (B), ALS patient using a cane (C), and ALS patient using a walker (D). The patient using a cane reported they are "turning alone or adjusting sheets but with great difficulty" on ALSFRS—R Q7, "walking with assistance" on Q8, and "needing assistance" on Q9. The patient using a walker reported they are "somewhat slow and clumsy but do not need help" on Q7, "walking with assistance" on Q8, and "unable to do" on Q9.
